# Risks of Subsequent Primary Extracolonic Cancers for Colorectal Cancer Survivors: A Study Protocol for the Development and Validation of a Risk Prediction Model

**DOI:** 10.64898/2025.12.08.25341861

**Authors:** Ye Kyaw Aung, Mark A. Jenkins, Nancy N. Baxter, Aung Ko Win

## Abstract

**Background:** The risk of developing a second cancer following colorectal cancer poses a significant challenge for colorectal cancer survivors, as 10–20% of survivors experience a subsequent primary cancer. These additional diagnoses are significantly associated with lower survival rates and increased morbidity compared with those who do not develop a subsequent primary cancer. Identifying survivors at the highest risk for subsequent primary cancers is imperative for preventive strategies and surveillance. This study protocol outlines the development and internal and external validation of risk prediction models, estimating individual risks of subsequent extracolonic cancer (cancers outside the colon and rectum) over 3-, 5-, 10-, and 15-year periods.

**Methods:** This study will include adult patients aged 18 years or older with a history of invasive colon, rectal, or colorectal cancer diagnosed within one year prior to recruitment; at least one year of follow-up post-recruitment; no prior nonskin cancers; and no genetic predispositions, such as Lynch syndrome. Data will be sourced from the Colon Cancer Family Registry Cohort, which recruits participants from population-based cancer registries. The primary outcome is the first diagnosis of a subsequent primary extracolonic cancer, excluding metastases or recurrence of any primary cancer. Candidate predictors will include sociodemographic factors, comorbidities, lifestyle factors, clinicopathological factors, and hormonal factors such as menopausal status and hormone use (in women).

Initial predictor selection will be performed using five methods: backward stepwise elimination via flexible parametric regression, Cox regression, Fine and Gray subdistribution hazard regression, Lasso Cox regression, and elastic net regression. Predictive performance will be assessed through accuracy measures, discrimination (C and D statistics), R^2^ statistics, and calibration plots to select the best-performing final model. Internal validation will involve bootstrapping 500 samples to estimate optimism-corrected C statistics. We will calculate individualized risks for subsequent extracolonic cancer at 3-, 5-, 10-, and 15-year intervals using shrinkage-adjusted beta coefficients of the selected predictors in the final model, along with baseline hazards derived from two approaches—one that accounts for death as a competing risk, and one that does not. External validation will be performed by testing the final model on an independent cohort from the Melbourne Collaborative Cohort Study.

**Discussion:** This model may help clinicians identify colorectal cancer survivors at increased risk of subsequent extracolonic cancers, enabling early detection, lifestyle modifications, and personalized screening strategies. Early intervention could subsequently reduce morbidity and improve long-term outcomes for these colorectal cancer survivors.

## INTRODUCTION

Colorectal cancer is the third most commonly diagnosed cancer in the world and the second leading cause of cancer-related death. In 2020, it was estimated that there were 1.93 million new cases and 916,000 deaths worldwide [1]. The implementation of early colorectal cancer screening, advanced treatments, and cancer surveillance strategies in developed nations [2–6] has significantly improved the overall 5-year survival rate. In the United States, for example, the survival rate increased from 59.2% in 1985 to 67.9% in 2015 [7], and in Australia, it rose from 54% in 1989–1993 to 71% in 2014–2018 [8].

This increased survival extends the period during which colorectal cancer patients are at risk of developing other primary cancers, which are not recurrences or metastases of the initial cancer. Prior research indicates that 10–20% of these survivors will be diagnosed with another primary cancer after their colorectal cancer diagnosis [9–18]. These subsequent primary cancers contribute significantly to poor survival and increased morbidity among these patients compared with those who do not develop another primary cancer [11, 12, 17, 18].

In an attempt to estimate this level of risk and risk factors, studies have compared the risk of subsequent cancer for those with a prior diagnosis of colorectal cancer to the risk of primary cancer for those in the general population (as a proxy for those without a prior diagnosis of colorectal cancer) [19–21] and identified factors contributing to the development of subsequent primary cancers in colorectal cancer survivors [11, 12, 22–26]. Some studies have additionally estimated the cancer risks for specific sites, such as subsequent primary cancers in the lung, breast, bladder, and prostate, along with their risk factors, including radiotherapy and lifestyle factors [26–29].

To date, three prognostic studies have outlined the characteristics of colorectal cancer patients with subsequent primary cancer that indicate an elevated risk of developing subsequent primary cancers. Using a population-based cohort from the Surveillance, Epidemiology, and End Results (SEER) study within the U.S. population, the first study estimated the overall high risk of developing any subsequent primary cancer [11]. Two more recent studies estimated these risks separately for male and female patients and estimated the survival rates and cumulative incidence of the five most commonly diagnosed subsequent primary cancers [12, 30].

The main limitation of these published studies, however, is that they do not include well-known cancer risk factors, such as treatment for the initial primary cancer, family history, Lynch syndrome, lifestyle factors, and pathological characteristics of the first colorectal cancer. In addition, they include primary cancers diagnosed within 12 months following the first colorectal cancer and therefore were subject to misclassification of the outcomes, as some of these could have been metastases [11, 12, 30].

To overcome these deficits, our study will consider all well-known risk factors for primary cancer when developing the prognostic model and define the outcome as a primary cancer reported at least one year after the initial diagnosis of colorectal cancer. We will limit the assessment to the occurrence of subsequent primary cancers at extracolonic sites. Moreover, we will include only colorectal cancer patients without underlying genetic manifestations given that carriers of DNA mismatch repair (MMR) gene mutations have a known high risk of developing a subsequent primary cancer [31].

The primary objective of this study is to develop a risk prediction model to identify which colorectal cancer survivors are at the highest risk of developing subsequent primary cancers at extracolonic sites by using a large international population-based study, the Colon Cancer Family Registry Cohort (CCFRC) [32, 33]. We subsequently aim to create a clinical assessment tool utilizing the coefficient values of the selected predictors in the final model. This innovative tool will quantify 3-, 5-, 10-, and 15-year individualized risks of developing a subsequent primary cancer at extracolonic sites (cancers other than colorectal cancer) for both male and female colorectal cancer survivors. Moreover, the developed risk prediction model will undergo internal validation through bootstrapping of the original dataset and external validation by assessing its performance and clinical utility in an independent large cohort study from the Melbourne Collaborative Cohort Study (MCCS) [34].

## METHODS AND ANALYSIS

This study protocol is reported in alignment with the Transparent reporting of a multivariable prediction model for individual prognosis or diagnosis (TRIPOD) statement (*Supplemental Table 1*) [35, 36].

### Source of data

For this study, we will use data from the CCFRC for model development (training) and internal validation and the MCCS for external validation.

The CCFRC consists of 42,489 study participants from 15,049 families recruited between 1998 and 2012. The study participants were from the United States of America, Canada, Australia, and New Zealand and included colorectal cancer patients from population-based cancer registries, non-colorectal cancer participants from population-based sources, and colorectal cancer patients from family cancer clinics and their relatives. Detailed personal and family history of cancer, lifestyle factors, and other information were collected at baseline, with follow-up surveys conducted every 4–5 years for updated information and vital status. Blood samples, tumor specimens, and clinical information were also collected. The average follow-up period was 9.1 years, with a total of 276,762 person-years for participants who completed a follow-up questionnaire. The follow-up rate among participants who completed the previous follow-up and who were still alive was 83% at the 5-year follow-up and 95% at the 15-year follow-up [32]. The 3^rd^ edition of the International Classification of Diseases for Oncology (ICD-O-3) was applied in coding primary cancer sites and their histology. Primary cancers were confirmed through pathology reports, medical records, corroboration by relatives, cancer registry reports, and/or death certificates, when available. Participants were systematically tested for germline mutations in colorectal cancer-predisposing genes. Details of the cohort profile have been reported by Jenkins et al., 2018 [32] and Newcomb et al., 2007 [33].

### Study participants

The study participants are adult patients aged at least 18 years who had a previous history of invasive cancer of the colon or rectum or colorectal cancer within one year prior to recruitment (the initial colorectal cancer diagnosis). Colon cancer is defined as any diagnosis of cancer within the proximal colon (C18.0, C18.2–C18.4), distal colon (C18.5–C18.7), or an unspecified site of the colon (C18.8, C18.9, and C26.0). Rectal cancer is defined as cancer of the rectosigmoid junction (C19.9) or rectum (C20.9 and C21.8). The appendix cancer (C18.1) was excluded. We will include participants diagnosed with initial colorectal cancer at the time of recruitment and identified only from population-based cancer registries in the United States, Canada, and Australia while clinic-based data were available only to New Zealand (Supplemental Figure 1). As colorectal cancer patients with inherited genetic colorectal cancer syndromes have specific guidelines for cancer surveillance and are known to be at high risk of subsequent cancer, we will exclude those with a pathologic or unclassified germline variant in the MLH1, MSH2, MSH6, PMS2, MUTYH, or EPCAM genes from our study. In addition, we will exclude colorectal cancer patients who had a diagnosis of any cancer (including melanomatous skin cancer) at least one year before being recruited for the study. Patients who were diagnosed with two or more cancers (other than melanomatous skin cancer, synchronous colorectal cancer, or appendix cancer) within one year of the initial colorectal cancer diagnosis (synchronous extracolonic cancers) will also be excluded. The included cases must have a follow-up period of at least one year after the first primary colorectal cancer diagnosis to reduce the likelihood of misclassifying a metastatic or spread of colorectal cancer as a subsequent primary extracolonic cancer (Supplemental Figure 2). However, those patients diagnosed with another colorectal cancer within one year of the initial cancer diagnosis will be included but flagged as having a history of synchronous colorectal cancer diagnosis (Supplemental Figure 1). The observation time ranges from the year of initial colorectal cancer diagnosis until the earliest year of subsequent cancer diagnosis, the year of death, or the year of last follow-up, whichever occurs first.

### Primary outcome

The primary outcome of this study will be the first recorded diagnosis of a primary extracolonic cancer, defined as cancer in solid or nonsolid organs other than the colon or rectum, which is diagnosed after the age at diagnosis of the first colorectal cancer and is neither a metastasis nor a recurrence from any primary cancer (C00–C96, excluding C43–C44 and C18–C20). Subsequent cancers in extracolonic sites reported at least one year after the initial colorectal cancer diagnosis date will be termed new primary cancer (subsequent cancer), whereas cancers reported within one year of the initial (colorectal) cancer diagnosis will be termed synchronous, which will be subsequently excluded on the basis of the criteria mentioned above.

### Candidate predictors

This cohort dataset includes information on cancer status; self-reported participants’ demographics; lifestyle factors; family history; dietary factors; anthropometric measurements; history of non-communicable diseases; medications; and women’s reproductive history and hormone use (Table 1). All available data collected at baseline were included only for selecting candidate predictors during model development. Based on the available data, the two approaches will be used to ensure that clinically relevant predictors and known risk factors associated with subsequent primary cancers are included in the model development: (1) incorporating risk factors previously reported in the literature to be associated with subsequent cancer; and (2) including predictors deemed clinically relevant based on expert advice from clinicians.

**Table 1.**
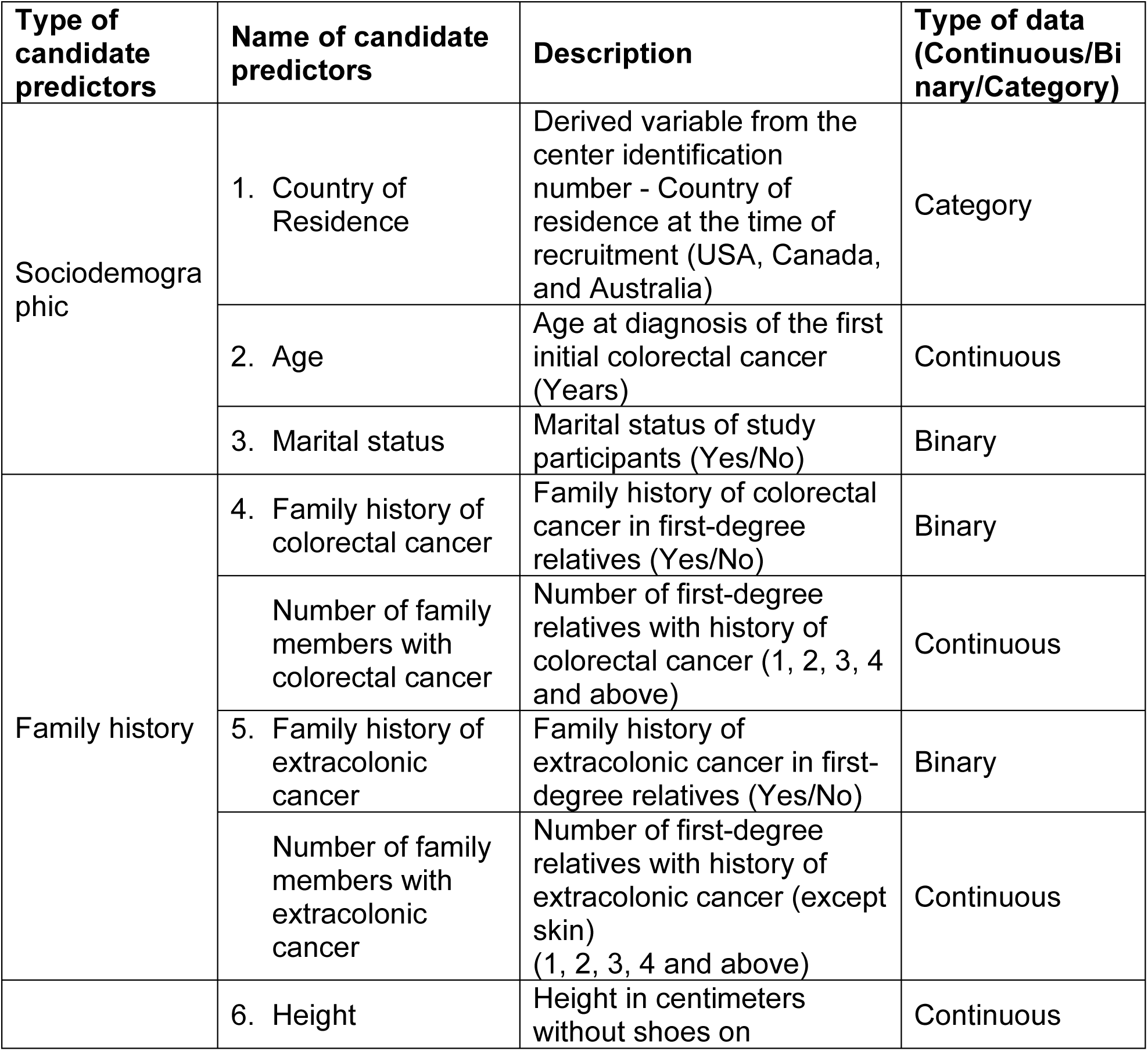
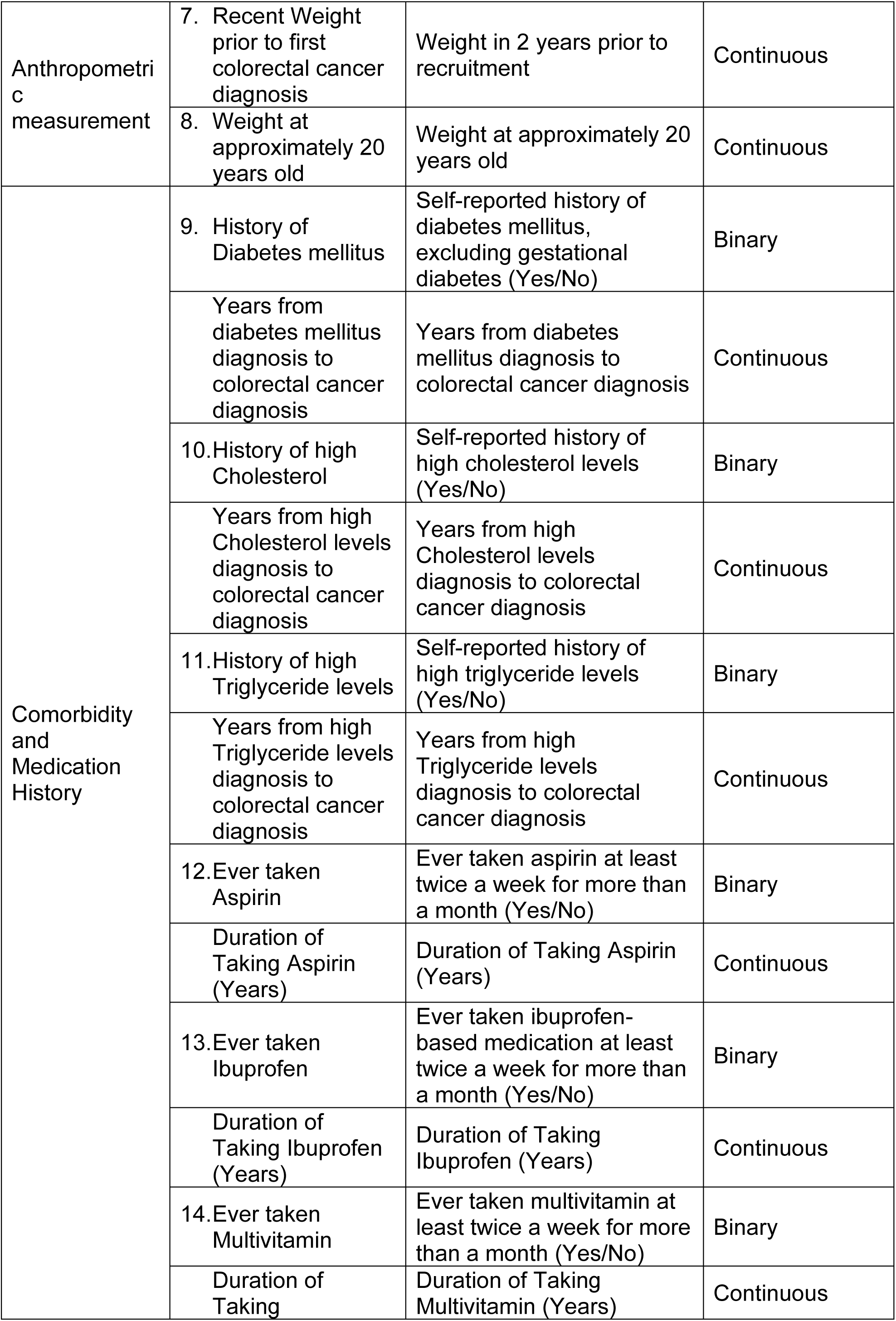
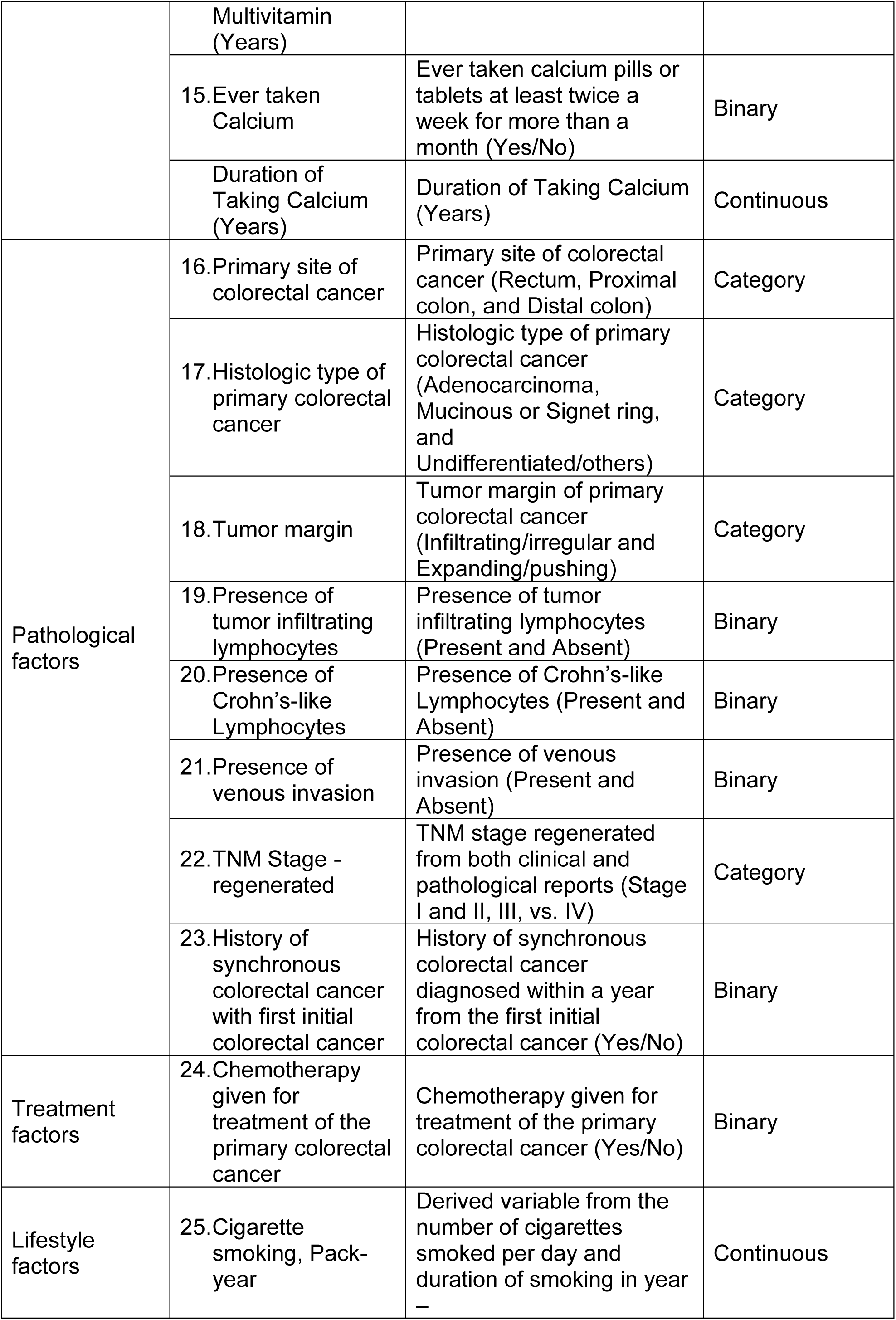
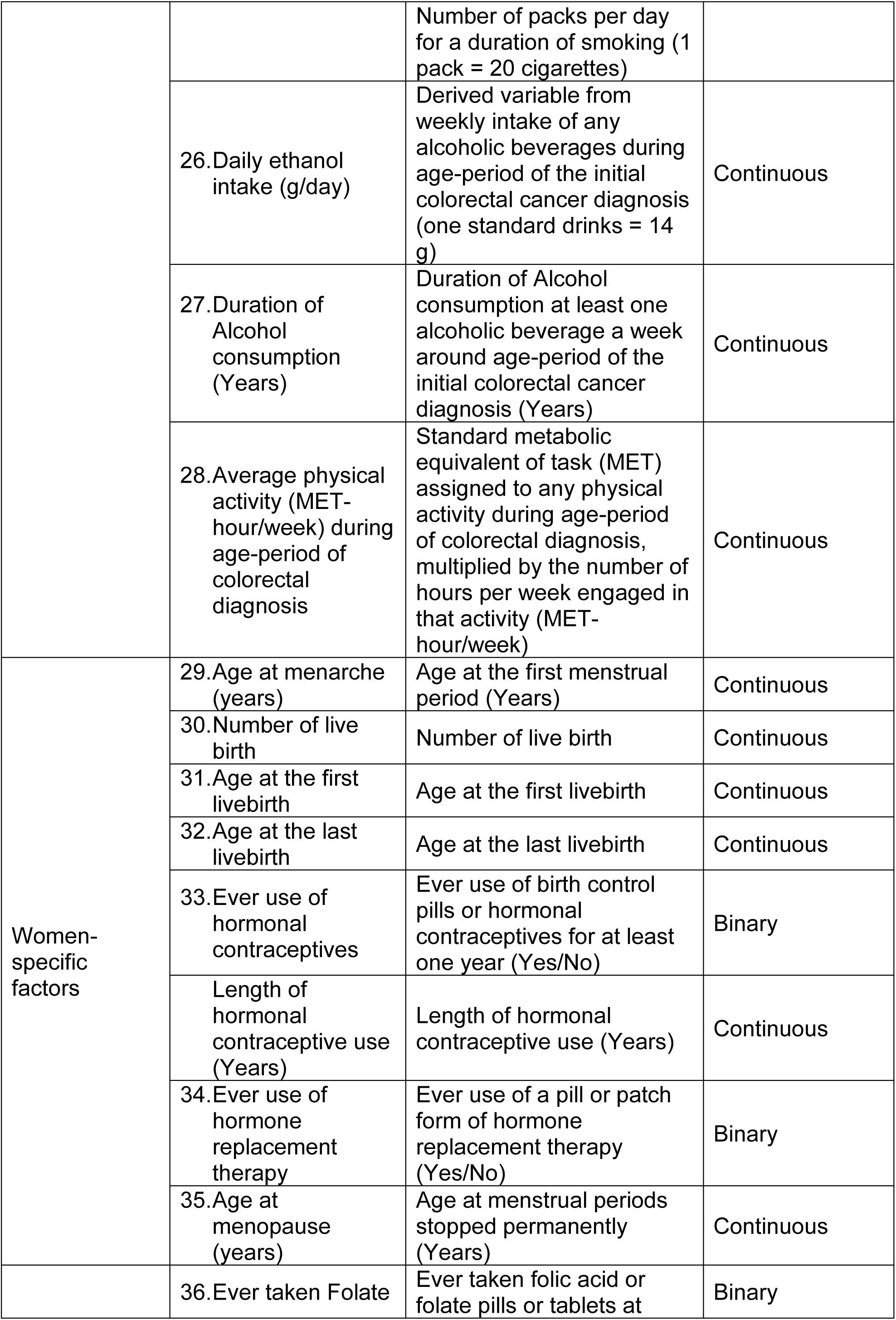
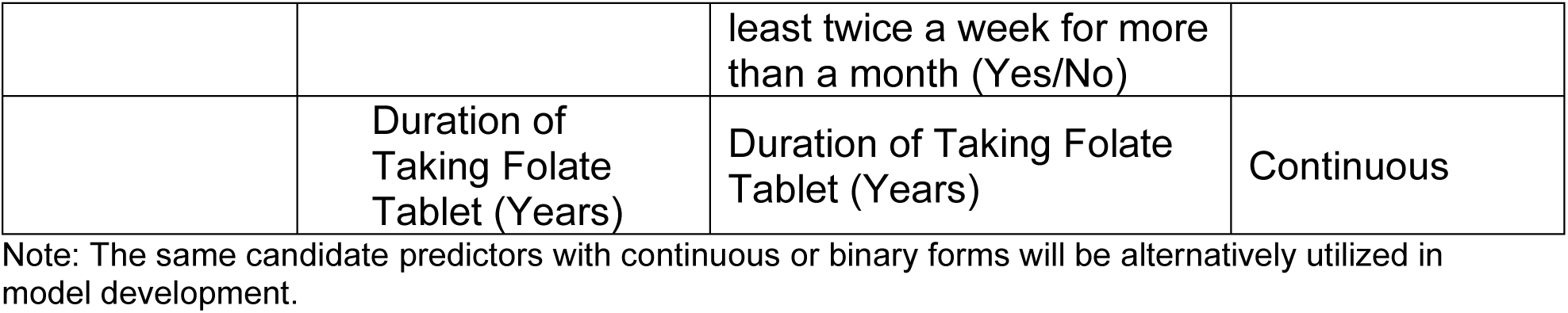
Pre-specified candidate predictors for subsequent cancer risk.

### Sample size

Regarding the sample size, we will determine the power by calculating the number of events required for the candidate predictors considered in the model, which is called “events per parameter” (EPP). The EPP can be described using the following mathematical equation:

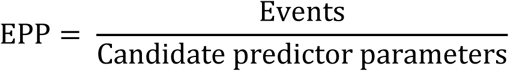

where Events= the number of the outcome primary cancers at extracolonic sites diagnosed after colorectal cancer [37].

Alternatively, the STATA command “pmsampsize” will be applied to determine the EPP [38]. We will pre-specify the number of predictor parameters included in the model (maximum 41 parameters), the desired heuristic shrinkage factor (assumed to be 0.9), the overall risk of subsequent primary cancer in the colorectal cancer patients (assumed to be 10%), and the expected value of the Cox-Snell R-square of the developed model based on previous evidence (otherwise assumed to be 0.15). The calculation yields a minimum sample size of 2194, which is required for developing a risk prediction model, with an EPP value of 5.0. If the EPP value is less than five, we adjust the number of predictor parameters on the basis of the two approaches mentioned above. This sample size calculation will be applied to both male-specific and female-specific models.

### Statistical analysis

#### Step 1: Descriptive analysis

The characteristics of colorectal cancer patients with and without subsequent extracolonic cancer will be summarized and presented in terms of numbers and percentages. Continuous predictors will be retained as continuous predictors during model development, except in situations of skewed distributions or nonlinear associations with the outcome. Dichotomizing or categorizing continuous predictors is, however, deemed unnecessary, inefficient, biologically implausible, and therefore not recommended for prognostic model development [39]. In cases where predictors display non-normal distributions and/or nonlinear distributions with the outcome of interest, simple transformations (e.g., natural logarithms), restricted cubic splines (with considerations of the number of knots), or fractional polynomials will be employed. If there are multiple continuous predictors to be tested for both distributions and linear associations, the multiple fractional polynomial process will be utilized to access the best-fit model, determining whether nonlinear continuous predictors should be included or transformed to minimize deviance. This process will be conducted in STATA via the “mfp” command while maintaining a nominal type 1 error with an alpha value of 0.05 [40].

#### Step 2. Handling missing data

Regarding missing data, we will examine whether systematic differences exist between missing and observed values among patients with or without subsequent primary cancer. If systematic missing is detected and a variable has more than 25% missing data, we will consider excluding it from the analysis. Otherwise, missing values will be addressed using the missForest multiple imputation method–a random forest-based algorithm implemented in the missForest package in R programming software. This missForest approach consistently provided lower imputation errors compared to other common approaches, including MICE, particularly when dealing with mixed data types and datasets with intricate structures [41–43]. If some previously identified predictors–such as certain pathological features of colorectal cancer–have up to 60–70% missingness, we will initially include these variables in model development. At the same time, we will conduct multiple imputations and develop models both with and without including variables with high proportions of missing data, to assess their impact on model performance.

#### Step 3. Exploratory univariable analysis

The cumulative risk of developing subsequent primary cancer will be visualized via the Nelson–Aalen cumulative hazard function and smoothed hazard estimates [44]. To estimate the risk of subsequent extracolonic cancers, we will consider the observation time from the year of initial colorectal cancer diagnosis until the year of death, the earliest year of subsequently diagnosed cancer, or the year of last follow-up, whichever occurs first. Additionally, preliminary univariable associations of candidate predictors with the outcome of subsequent primary cancer will be tested and measured via a univariable flexible parametric model [45] and Cox regression analysis [44].

#### Step 4. Collinearity

Pearson’s correlation test will be utilized for continuous predictors, whereas Pearson’s chi-square test [44] will be applied for categorical predictors to assess the relationships between candidate predictors. When significant correlations are observed between predictors (the correlation coefficient is assumed to be >0.5), only one of the correlated predictors will be included in the model development via the following approach. Both terms for highly correlated predictors will be jointly tested and either included or excluded by placing each term in brackets in the multivariable model selection process.

#### Step 5. Risk prediction model development Flexible parametric model

Initially, a multivariable flexible parametric model will be employed to construct a full model, encompassing all candidate predictors. We will subsequently conduct backward stepwise elimination, eliminating predictors with a p value exceeding 0.1 in order of their p value (largest first). Each excluded predictor will then be reintroduced back into the model one at a time to assess whether it should be reinstated, ensuring that no predictors meet the criteria for inclusion or exclusion. However, predictors deemed clinically significant on the basis of previous literature and the advice of clinical experts will be considered for re-inclusion in the model. The following inclusion criteria will additionally guide the addition and/or removal of predictors for choosing the final model:

- Wald test p value less than 0.1 for each predictor
- Log-likelihood test p value less than 0.05 for comparing models with or without a predictor.
- Akaike’s information criterion (AIC) setting up a p-value of 0.157 as a proxy, as suggested by Sauerbrei_1999 [46].

Modeling will be conducted via the STATA “stpm2” command for the multivariable flexible parametric model [45]. In situations involving multiple non-linear continuous predictors, we will adopt the “mfp” stacking approach [40].

### Cox regression model

A multivariable Cox regression model will be employed to construct a full model, encompassing all candidate predictors. With the STATA ‘swaic’ command, backward elimination from the full model will be implemented to select a set of predictors with the best Akaike information criterion [46]. During the initial development of the full Cox model, continuous candidate predictors for variables with non-normal distribution or non-linearity will be transformed as in the flexible parametric model, ensuring the consistent inclusion of candidate predictors.

### Fine and Gray subdistribution hazard regression

To account for death as a competing risk, Fine and Gray subdistribution hazard regression model [47] will be used to construct a full model encompassing all candidate predictors, ensuring consistent inclusion as in the flexible parametric model. Using the ‘stcrreg’ command in STATA [48], we will subsequently perform backward stepwise elimination by sequentially removing predictors with p-values exceeding 0.1, starting with the highest p-value. As mentioned above in the flexible parametric model, we will employ the consistent approach to confirm that no predictors meet criteria for further inclusion or exclusion.

Furthermore, we will assess non-proportional hazards by examining interactions between the hazard ratio for each selected predictor from the final model and time in STATA, utilizing the ‘tvc’ option. Additionally, interaction terms between the selected predictors will be included in the final model, particularly to account for potential interactions among these predictors [45]. Finally, we will conduct stratified analysis on the basis of sex, and female-specific candidate predictors will be included in the development of the prognostic model for female patients.

### Uniform shrinkage

If the final model requires adjustment for optimism, the uniform shrinkage (S) method will be employed. This involves multiplying the beta coefficients of the linear predictors by the S value and re-estimating the baseline hazard at each time point. The estimate of S will be obtained from a heuristic shrinkage factor estimate [49].

#### Step 6: Model development via modern approaches (Lasso Cox & Elastic Net Methods)

As alternative methods for model development that consider both model estimation and shrinkage simultaneously, we will utilize the least absolute shrinkage and selection operator (LASSO) [50] and elastic net [51]. These methods facilitate predictor selection and penalization concurrently by shrinking each predictor effect toward zero. The “lasso cox” command in STATA 18 enables implementation of the former approach, whereas the “elasticnet cox” command is suitable for the latter. Before proceeding with model development, we will first assess whether the sample size is appropriate by checking the required minimum sample size (2194) and EPP value (5.0), as calculated above.

#### Step 7. Model performance

We will test model performance by measuring calibration and discrimination.

### Calibration

Calibration will be calculated as the ratio of expected to observed event probabilities calculated for all individuals and within risk subgroups at one or more time points via the following equation: [45]

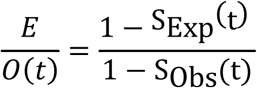

In this equation, E represents the expected number of subsequent extracolonic cancers, O refers to the observed number of subsequent extracolonic cancers, and t refers to time, whereas S_Exp_(t) and S_Obs_(t) presents the survival probability predicted by the model at time t, and the actual survival probability estimated from the observed data at time t, respectively.

A calibration slope will be generated by fitting the model with the linear predictor as the predictor shown in the formula below.

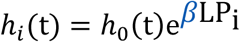

In this equation, hᵢ(t) represents the risk (hazard) of the event occurring at time t for individual ‘*i’*, given their covariates; h_0_(t) is the hazard function at time t when all covariates are at their reference (or baseline) values; LP_i_ is a linear predictor for individual ‘i’; and e is the exponential function. Additionally, β represents the calibration slope, where a value of β less than 1 indicates model overfitting, whereas a value of β greater than 1 indicates model underfitting. The calibration slope represents the slope of the regression of observed survival outcomes on the predicted prognostic index. A value of β close to 1 suggests that the prediction model is well calibrated [45, 52, 53].

### Discrimination

We will assess discrimination via Royston and Sauerbrei’s D statistic, a discrimination measure that quantifies the observed separation between observations with low and high predicted risk. This statistic allows us to interpret the log hazard ratio by comparing two equal-sized risk groups defined by dichotomizing the distribution of the patient prognostic indices at the median value. We will use the STATA package “str2d” to produce D statistics as well as R^2^_D_, a measure of prognostic separation ranging between 0 and 1 [53, 54].

Additionally, we will utilize Harrell’s C_H_ concordance measure (the C statistic), which represents the probability that, of a randomly selected pair of patients, a patient with a shorter survival time (corresponding to an event) has a higher predicted risk. This measure ranges between 0.5 and 1, where a value of 0.5 indicates no discrimination and a value of 1 indicates perfect discrimination [53, 55].

Furthermore, using the fully prognostic model, we will compute model-based mean survival curves for four distinct risk groups, defined by categorizing the prognostic index at the 16^th^, 50^th^, and 84^th^ centiles. These cutoff centiles are recommended by the Cox method to minimize the loss of information when discretizing a normally distributed continuous variable into a given number of groups [56, 57]. We will then plot Nelson–Aalen cumulative hazard function curves [44, 45] to assess whether clear separation exits between the predicted risk groups. Additionally, we will compare the observed (Kaplan-Meier) and predicted mean survival curves for each group to evaluate the conceptual validity of the prognostic model.

#### Step 8. Internal validation

Bootstrapping will be utilized to assess the internal validity of the model [58]. If multiple imputations for missing values are deemed necessary, the following approaches will be applied to determine the best-optimized model. Initially, multiple imputations will be performed, followed by bootstrapping with replacement of the imputed data, creating 500 datasets of the original sample size. Automatic predictor selection will be simultaneously executed across the bootstrapped datasets via the backward stepwise selection approach, resulting in a single model. The performance measures will be calculated for each of the 500 datasets and then averaged across all the bootstrapped datasets.

Finally, we will compare five different models—backward stepwise elimination via flexible parametric regression; backward stepwise elimination using Cox regression; backward stepwise elimination using Fine and Gray subdistribution hazard regression; Lasso Cox regression; and elastic net regression—using the performance measures of both developed models and internal validations, such as C statistics, D statistics, R^2^ statistics, and calibration plots. Ultimately, the final model demonstrating superior performance in predicting the outcome of interest and the inclusion of fewer predictors will be selected. We will report the coefficient values of the selected predictors in the final model and the baseline hazards for each model to create a sex-specific clinical assessment tool for estimating individual risks. Finally, we will estimate individualized risks for subsequent extracolonic cancer at 3-, 5-, 10-, and 15-year intervals using shrinkage-adjusted beta coefficients of selected predictors in the final model, along with baseline hazards derived from two approaches—models that account for death as a competing risk and models that do not. For the competing risks analyses, we will specifically employ sex-specific competing risk models by combining selected predictors with the Fine and Gray subdistribution hazard approach. This method directly models the cumulative incidence functions, providing accurate estimation of the probability of the occurrence of a subsequent extracolonic cancer while appropriately accounting for competing mortality risk [47].

#### Step 9. External validation

The final internally validated risk prediction model will undergo external validation by testing it against an independent cohort of individuals who have colorectal cancer from the Melbourne Collaborative Cohort Study, also known as Health 2020. This cohort comprised 41,513 participants (24,469 women and 17,044 men) residing in the Melbourne metropolitan area, predominantly aged between 40 and 69 years. Its primary aim was to prospectively investigate the roles of diet and lifestyle in the causation of cancer and other non-communicable diseases. Recruitment commenced between 1990 and 1994, with participants being actively followed up twice. The first wave of active follow-up occurred between 1995 and 1998, approximately 4 years after recruitment, and the second wave took place between 2003 and 2007. The former included a total of 36,335 (88%) of the original participants, with 778 (1.87%) having died before being contacted, whereas the latter comprised 28,240 (68%) of the original participants, with 3,007 (7.24%) deceased before the second follow-up. Additionally, passive follow-up occurred every 6 months to update participant addresses and identify incident cancer diagnoses and deaths, utilizing information from the Victorian Electoral Enrollment Register and through record linkage to the Victorian Cancer Registry (VCR) and the Victorian Registry of Births, Deaths, and Marriages. This cohort contains extensive quantitative data on cancer status, dietary and non-dietary factors, anthropometric measurements, history of non-communicable diseases, medications, reproductive history, and hormone use. RL Milne et al. thoroughly described and published this cohort in 2017 [34].

The study participants will include adult colorectal survivors aged at least 18 years, with a history of invasive colorectal cancer as their first primary cancer at the time of recruitment and no history of pathologic or unclassified variants in the MLH1, MSH2, MSH6, PMS2, MUTYH, or EPCAM genes. Additionally, we will apply the same inclusion and exclusion criteria as the CCFRC dataset to ensure consistency and comparability among study participants. The fully specified final model will be applied to this MCCS cohort to estimate the 3-, 5-, 10-, and 20-year risks of subsequent primary cancer in this external population. The linear predictor for the validated dataset will be estimated via regression coefficients, and the predictors of interest will be identified from the fully specified model. Subsequently, predictive model performance will be assessed by estimating discrimination through D statistics and calibration plots, following the methods mentioned above. In cases of systematic overestimation or underestimation of risk, the recalibration method will be applied. This involves adjusting the baseline hazard (intercept) by fitting a model that includes the linear predictor part of the risk score as a covariate and using the linear predictor as an offset term. This method allows us to adjust the model for the population of the validation data by re-estimating some of the model parameters and ensuring consistency in predictor weights within the score [59].

## LIMITATIONS AND STRENGTHS

While the CCFRC database is a large cohort of colorectal cancer patients, a significant limitation of this study lies in the presence of missing data, particularly concerning treatment and pathological factors. To mitigate this issue, we will only consider predictors with at most 25% missing data and employ multiple imputations, a widely accepted method for handling missing data, only after ensuring no systematic missing. However, if some previously identified predictors—such as certain pathological features of colorectal cancer—have up to 60–70% missingness, we will initially include them in model development while simultaneously conducting multiple imputations and developing models both with and without these high-missingness variables to assess their impact on model performance. Another limitation for the validation dataset (MCCS cohort) arises from certain predictors, such as a family history of cancer, which may not have been collected in the same manner as in the CCFRC study, although most candidate predictors were equally available in both the original and validated datasets. Third, the MCCS cohort will have a smaller number of study participants than the CCFRC cohort. In addition, we will include participants diagnosed with colorectal cancer who responded to questionnaires within 2 years of diagnosis, potentially reducing the number of study participants available for model validation. For a strength, colorectal cancer patients with underlying genetic manifestations, who may have a significantly greater risk of developing subsequent primary cancers, are not included in this study. As a result, the findings are more likely to be generalizable to colorectal cancer patients without underlying genetic diseases.

## CONCLUSION

Colorectal cancer survivors face a higher risk of developing subsequent primary cancers, leading to poorer prognoses and reduced survival rates. This study proposes a validated risk prediction model to help clinicians identify survivors at the highest risk of developing primary cancers beyond the colon and rectum. By identifying at-risk individuals, healthcare providers can effectively deploy targeted screening and lifestyle interventions. Ultimately, this model has the potential to serve as a crucial component in the personalized care of colorectal cancer survivors, not only improving survival rates but enhancing the overall quality of life for these individuals.

## Supporting information

Tripod Reporting Checklist

## Data Availability

The data will be available upon request from the data authories of the Colon Cancer Family Registry Cohort and the Melbourne Collaborative Cohort Study.

## DECLARATIONS

Ethics approval and consent to participate – The MCCS’s EC/EO reference number is EO2016/1/247 (valid until 01/04/2026) and the CCFRC’s approved project number is C-AU-1110-01 (valid until 31/12/2025). All the study participants or their legal representatives gave their consent to participate in the study. The study protocol was approved by the institutional human research ethics review boards at each center (The University of Melbourne’s Human Research Ethics Approval Number: 13094).

### Consent for publication

Not applicable.

### Availability of data and materials

The data will be available for researchers after obtaining approvals from the relevant authorities. https://coloncfr.org/for-researchers/collaborate-with-the-ccfr/ and https://www.cancervic.org.au/research/epidemiology/collaborations.

### Authors’ contributions

YKA, AKW and MAJ contributed to the conceptualization of the research. YKA developed the protocol and drafted the manuscript. AKW, MAJ, and NB provided close supervision to YKA and offered guidance on protocol development. All the authors critically reviewed the draft manuscript and approved the final version.

## Acknowledgments

YKA is supported by a postgraduate scholarship from the Melbourne Research Scholarship (Fee offset) at the University of Melbourne and a Graduate Research Studentship (Stipend) from the Australia Government’s National Health and Medical Research Council (NHMRC) Investigator Grants awarded to AKW. MAJ is an NHMRC Leadership Fellow (APP1195099). NB is a Deputy Executive Dean Faculty of Medicine and Health, University of Sydney. The authors would like to thank all study participants for their participation.

## Abbreviations

AIC: Akaike’s Information Criterion
CCFRC: The Colon Cancer Family Registry Cohort
EPP: Events Per Parameter”
LASSO: Least Absolute Shrinkage and Selection Operator
MCCS: The Melbourne Collaborative Cohort Study
MET: Metabolic Equivalent of Task
MMR: Mismatch repair gene
SEER: The Surveillance, Epidemiology, and End Results study
TRIPOD: The Transparent reporting of a multivariable prediction model for individual prognosis or diagnosis
VCR: The Victorian Cancer Registry

---

*Supplemental Figure 1. Inclusion Case scenario for any subsequent extracolonic primary cancer*

*Supplemental Figure 2. Exclusion Case scenario*

*Supplemental Table 1. TRIPOD Checklist: Prediction Model Development*

