## Supplemental Figures for "Risks of Subsequent Primary Extracolonic Cancers for Colorectal Cancer Survivors: A Study Protocol for the Development and Validation of a Risk Prediction Model"

### *S1. Inclusion Case scenario for any subsequent extracolonic primary cancer*

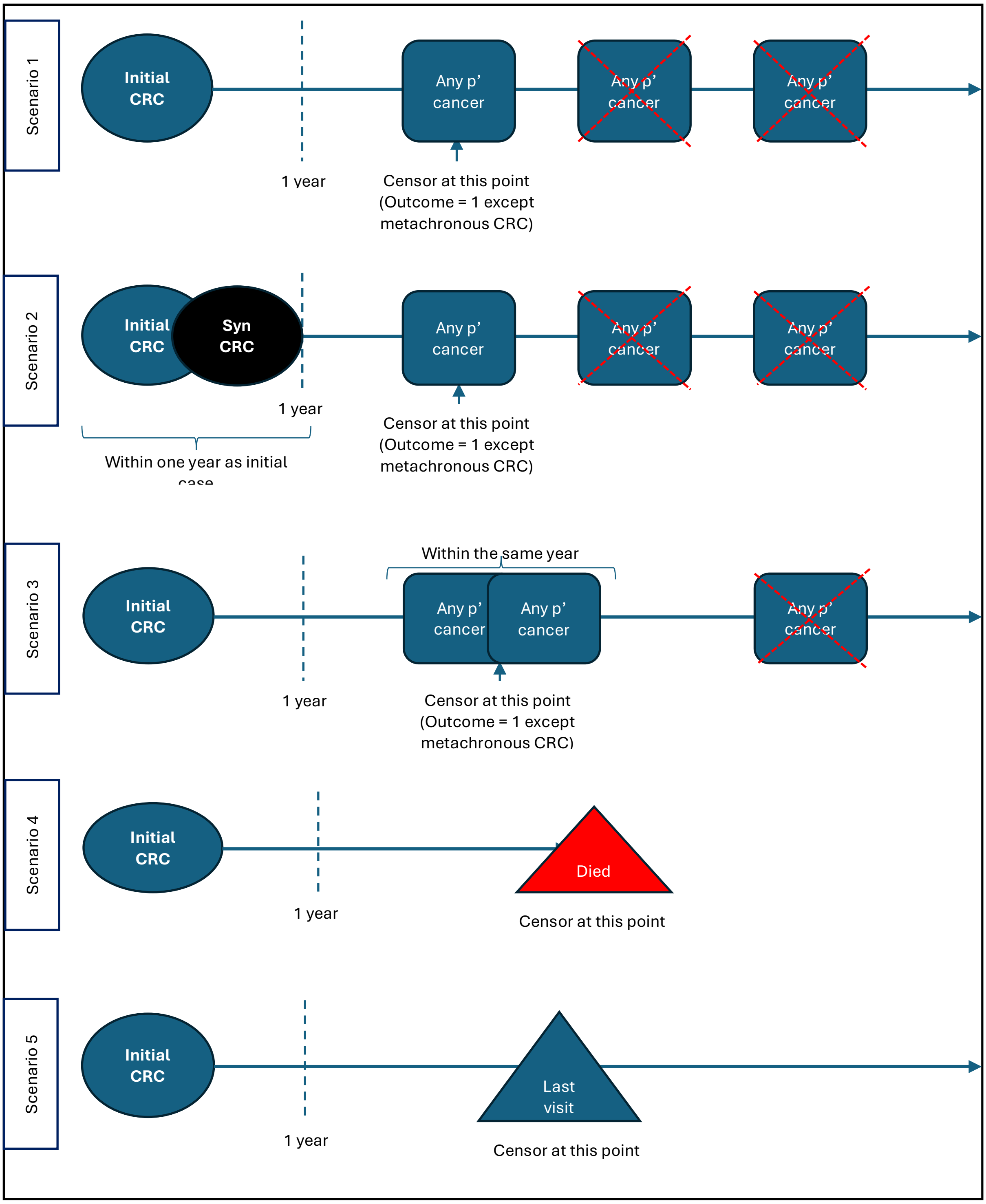

***S2. Exclusion Case scenario***

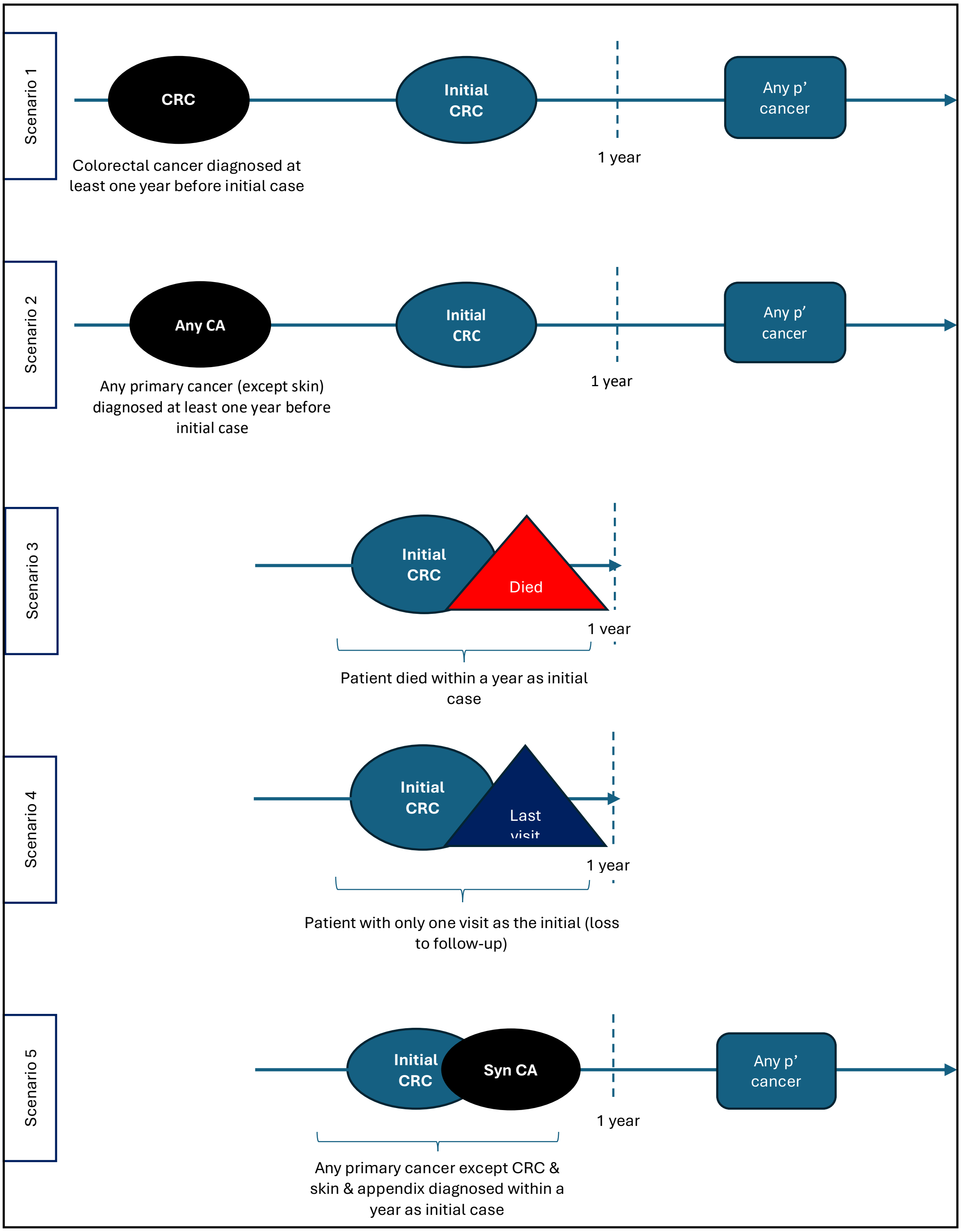
